# Phylogenetic estimates of SARS-CoV-2 introductions into Washington State

**DOI:** 10.1101/2021.04.05.21254924

**Authors:** Diana M. Tordoff, Alexander L. Greninger, Pavitra Roychoudhury, Lasata Shrestha, Hong Xie, Keith R. Jerome, Nathan Breit, Meei-Li Huang, Mike Famulare, Joshua T. Herbeck

## Abstract

**Background:** The first confirmed case of SARS-CoV-2 in North America was identified in Washington state on January 21, 2020. We aimed to quantify the number and temporal trends of out-of-state introductions of SARS-CoV-2 into Washington.

**Methods:** We conducted a phylogenetic analysis of 11,422 publicly available whole genome SARS-CoV-2 sequences from GISAID sampled between December 2019 and September 2020. We used maximum parsimony ancestral state reconstruction methods on time-calibrated phylogenies to enumerate introductions/exports, their likely geographic source (e.g. US, non-US, and between eastern and western Washington), and estimated date of introduction. To incorporate phylogenetic uncertainty into our estimates, we conducted 5,000 replicate analyses by generating 25 random time-stratified samples of non-Washington reference sequences, 20 random polytomy resolutions, and 10 random resolutions of the reconstructed ancestral state.

**Results:** We estimated a minimum 287 separate introductions (median, range 244-320) into Washington and 204 exported lineages (range 188-227) of SARS-CoV-2 out of Washington. Introductions began in mid-January and peaked on March 29, 2020. Lineages with the Spike D614G variant accounted for the majority (88%) of introductions. Overall, 61% (range 55-65%) of introductions into Washington likely originated from a source elsewhere within the US, while the remaining 39% (range 35-45%) likely originated from outside of the US. Intra-state transmission accounted for 65% and 28% of introductions into eastern and western Washington, respectively.

**Conclusions:** There is phylogenetic evidence that the SARS-CoV-2 epidemic in Washington is continually seeded by a large number of introductions, and that there was significant inter- and intra-state transmission. Due to incomplete sampling our data underestimate the true number of introductions.

---

The SARS-CoV-2 pandemic likely first emerged in China in late 2019, and by January 2021 there have been over 100 million confirmed cases and over two million deaths due to COVID-19 worldwide.^1^ The first confirmed SARS-CoV-2 infection in North America was identified in Washington State (WA), in January 2020. In February a second case was identified in WA,^2^ and preliminary genomic epidemiological analyses reported that this case belonged to the same transmission chain as the first case and suggested substantial cryptic transmission.^3^ While subsequent analyses showed that this second case was likely due to a separate introduction,^4^ the initial report spurred a rapid public health response that eventually included school closures and a general lockdown (“stay at home” measures).^5^

As of January 2021, there have been over 300,000 confirmed cases in WA and over 4,500 deaths due to COVID-19. Within WA the epidemic impact is geographically, demographically, and temporally heterogeneous; there has been substantial variation among counties and ZIP codes in confirmed case counts over time. In particular, eastern and western WA, which are separated by the Cascade mountain range, experienced distinct outbreaks over the spring and summer of 2020 (Figure 1A) and a large proportion of overall confirmed SARS-CoV-2 cases were reported in King and Yakima counties (Figure 1B). As in other locations in the United States (US) and globally, this temporal heterogeneity is likely partially explained by variation in the efficacy of non-pharmaceutical interventions and public health measures such as lockdowns and mask mandates.

**Figure 1.**
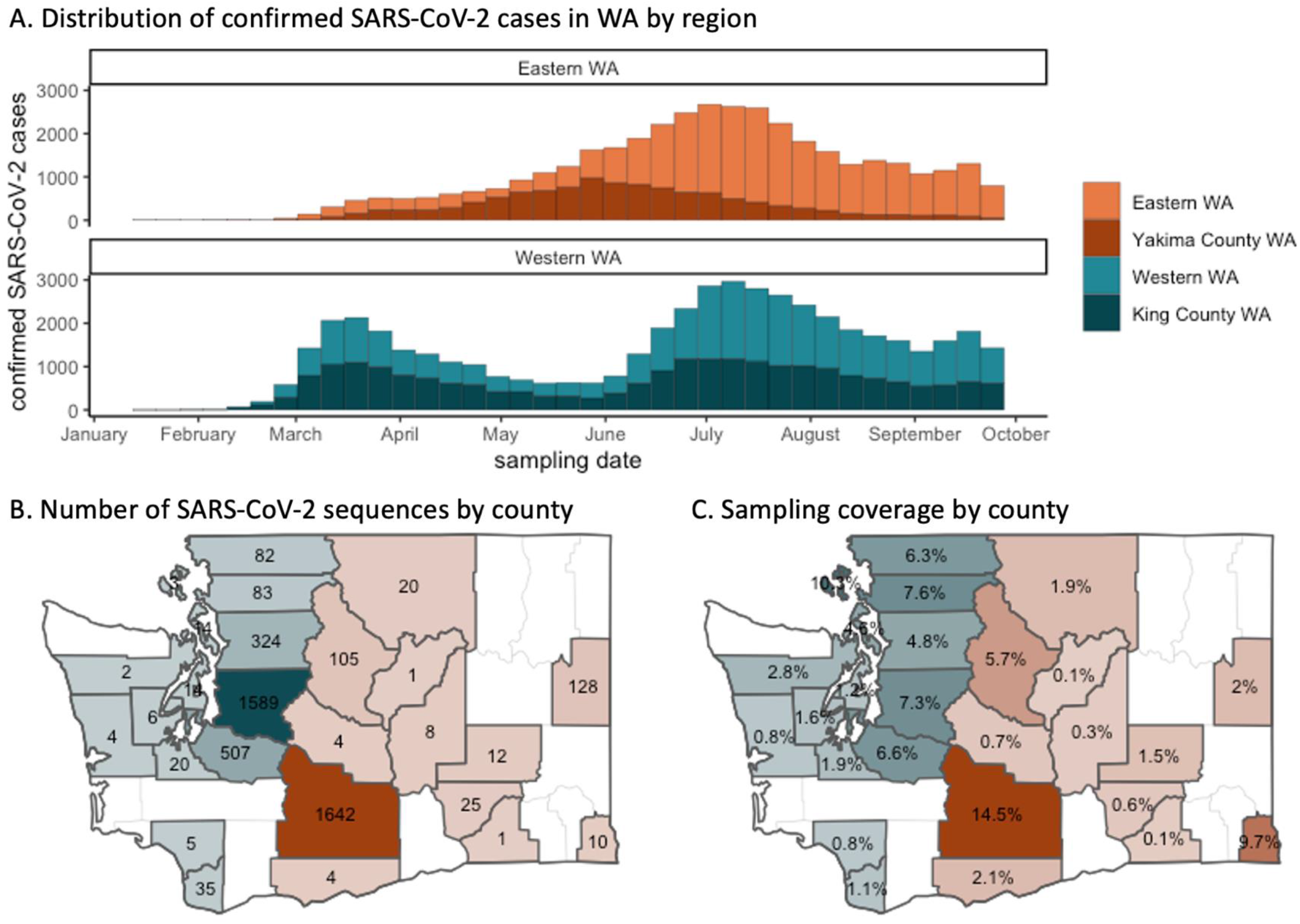
Geographic variation in confirmed SARS-CoV-2 cases, genome sequences, and sampling coverage in WA.

The SARS-CoV-2 pandemic has resulted in an unprecedented global scientific response, including the rapid sequencing and sharing of SARS-CoV-2 genomes; by October 2020, >120,000 SARS-CoV-2 genomes had been shared online. The analysis of this open data has provided many insights into the zoonotic origin and patterns of global spread of SARS-CoV-2, the development of vaccine candidates, and the surveillance and identification of novel genetic variants of potential public health interest.^4,6−10^ The present analysis aimed to combine publicly shared genomic data with limited linked clinical data in order to understand the role of introductions of SARS-CoV-2 during the initial outbreak and through the summer of 2020. Using phylogeographic methods, we aimed to quantify the number, timing, and likely geographic source of out-of-state introductions of SARS-CoV-2 and describe intra-state transmission patterns between eastern and western WA.

## METHODS

### Data Source

We used full genome SARS-CoV-2 nucleotide sequences obtained from the Global Initiative on Sharing Avian Influenza Data (GISAID, gisaid.org), collected between December 2019 and September 2020. We gratefully acknowledge the originating laboratories responsible for obtaining the specimens and the submitting laboratories where genetic sequence data were generated and shared via the GISAID Initiative, on which this research is based. Most SARS-CoV-2 sequences available from GISAID are linked to geographic data, including the region, country, and state from which the sequence was sampled. Among sequences sampled in WA over this timeframe, and available from GISAID, data on the county of residence was available for only 53% of sequences. To address this missing data, we obtained additional information on the county from which each sample was obtained from the University of Washington’s Virology Lab, which has performed the majority of SARS-CoV-2 full genome sequencing in WA state. Lastly, we used publicly available data on the weekly count of confirmed SARS-CoV-2 cases by county available through the WA Department of Health and estimates of daily SARS-CoV-2 incidence for WA from the Institute for Disease Modeling.^11,12^

### Sequence analyses

Prior to phylogenetic analyses, we identified and excluded SARS-CoV-2 genome sequences without a complete sampling date, that were incomplete (length <29kbp), or that were low quality (>500 Ns). We aligned sequences with MAFFT and identified SARS-CoV-2 lineages using the PANGOLIN (Phylogenetic Assignment of Named Global Outbreak LINeages, github.com/cov-lineages/pangolin) tool.^13,14^

Our analysis pipeline aimed to accommodate issues with SARS-CoV-2 genetic diversity (relative to the timeframes of transmission and accumulation of viral diversity) that lead to challenges with phylogenetic resolution. We generated a large number of replicate analyses to quantify the uncertainty in our estimates. First, from the full set of global sequences from GISAID (through September 2020) we generated 25 different samples that each included: all WA sequences (N=4918); the closest non-WA sequences for each WA sequence in our sample (based on raw genetic distance; N=5056); a random time-stratified sample of the remaining non-WA sequences (N=1447); and, the Wuhan/Hu-1/2019 reference sequence. Each sample was then stratified by PANGOLIN lineage (A, B.1, B.1.1, B.1.1.X, and B.X, for X≠1). Our choice of five lineages for stratification was based on the phylogeny of WA sequences, and we chose the sequence groups that aligned with monophyletic clades (Figure 2). There are several different nomenclatures for SARS-CoV-2 lineages, therefore, we include both the PANGOLIN and the corresponding GISAID lineages names.

**Figure 2.**
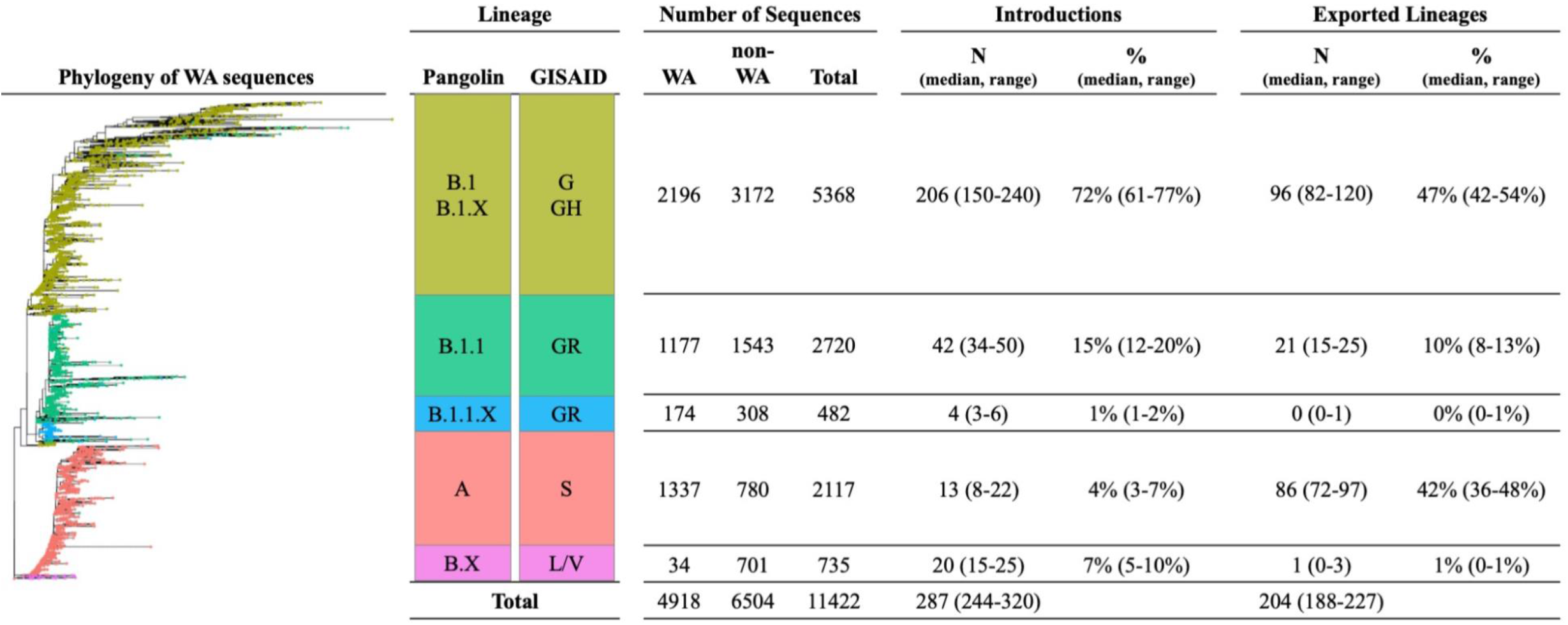
Estimates number and proportion of SARS-CoV-2 introductions and exports by lineage.

We then reconstructed phylogenies for each of the 25 samples stratified by lineage (N=125 trees) using IQTREE (HKY+G4 substitution model).^15^ Next, for each phylogeny we randomly resolved all polytomies with 20 random resolutions per phylogeny, which resulted in 500 bifurcating trees per lineage. Most of our SARS-CoV-2 phylogenies were characterized by a large number of polytomies; this step allowed us to estimate uncertainty in our estimates that may be due to poor phylogenetic resolution. Next, we time-calibrated each bifurcating tree using the *treedater* algorithm, assuming a strict molecular clock.^16^ We assumed a mean rate of evolution of 0.001 subs/site/year and constrained the rate to be between 0.0009 and 0.0011 subs/site/year. This allowed us to estimate to dates of each internal node and the time to most recent common ancestor (MRCA) for each tree.

Lastly, to estimate the number of WA import and export events, we used ancestral state reconstruction to reconstruct the likely state (WA or non-WA) of each node using maximum parsimony methods, implemented with the *phangorn* package in R. For each phylogeny (i.e. each polytomy-resolved replicate of each PANGOLIN lineage from the 25 sub-samples), we counted the number of introductions to WA and the size of the resulting WA subclades. We identified directional transmission events when the sequential inferred ancestral state (i.e. geographic sampling location) of the internal nodes were not identical when moving from the root of the tree toward the tip. WA subclades were defined as downstream clades that included only WA sequences. We estimated the date of each introduction into WA using the inferred date of the internal node date for the MRCA of each WA subclade. The same approach was used to count lineages exported from WA state, with the opposite direction for each reconstructed node state change.

We also applied ancestral state reconstruction to estimate the likely geographic source of each introduction into WA state (e.g. from outside of the US or from elsewhere within the US), as well as to estimate introductions that occurred within WA state, between eastern and western WA. For the latter of these two analyses, we excluded the 5.5% of WA sequences (N=270) for which the county of sampling was unknown. We included 10 random resolutions of each reconstructed geographic source to account for ambiguous reconstructions.

We provide summary statistics on sequence sampling coverage (overall and by WA county) using both confirmed cases and estimated incident cases as the denominator. Lastly, we estimated the ratio of the number of introductions (representing an individual who was infected outside of WA) to the total number of WA sequences as a proximal measure of the relative contribution of introductions versus local transmission. This approach allows us to directly compare (via this proxy estimate) our results to those of Müller et al, in which phylodynamic simulation methods estimated that between 1% and 10% of WA cases were imported.^17^ For each of our estimates, we report the median and range aggregated across the 5000 replicate analyses. All statistical analyses were conducted using R statistical software version 3.6.2. The use of residual clinical specimens for sequencing was approved by the institutional review board at the University of Washington with a waiver of informed consent.

## RESULTS

Each analytic sample (replicate) included 11,422 SARS-CoV-2 genomes: 4,918 WA sequences and 6,504 non-WA sequences (Figure 2). Overall, our analysis included sequences for 6% of all confirmed SARS-CoV-2 cases in WA state and 1.8% (95% CI: 1.2-2.8%) of estimated incident SARS-CoV-2 cases.^11,12^ The sample coverage for WA state varied modestly between March and June, and decreased significantly in July and late August (Figure 3A).

**Figure 3.**
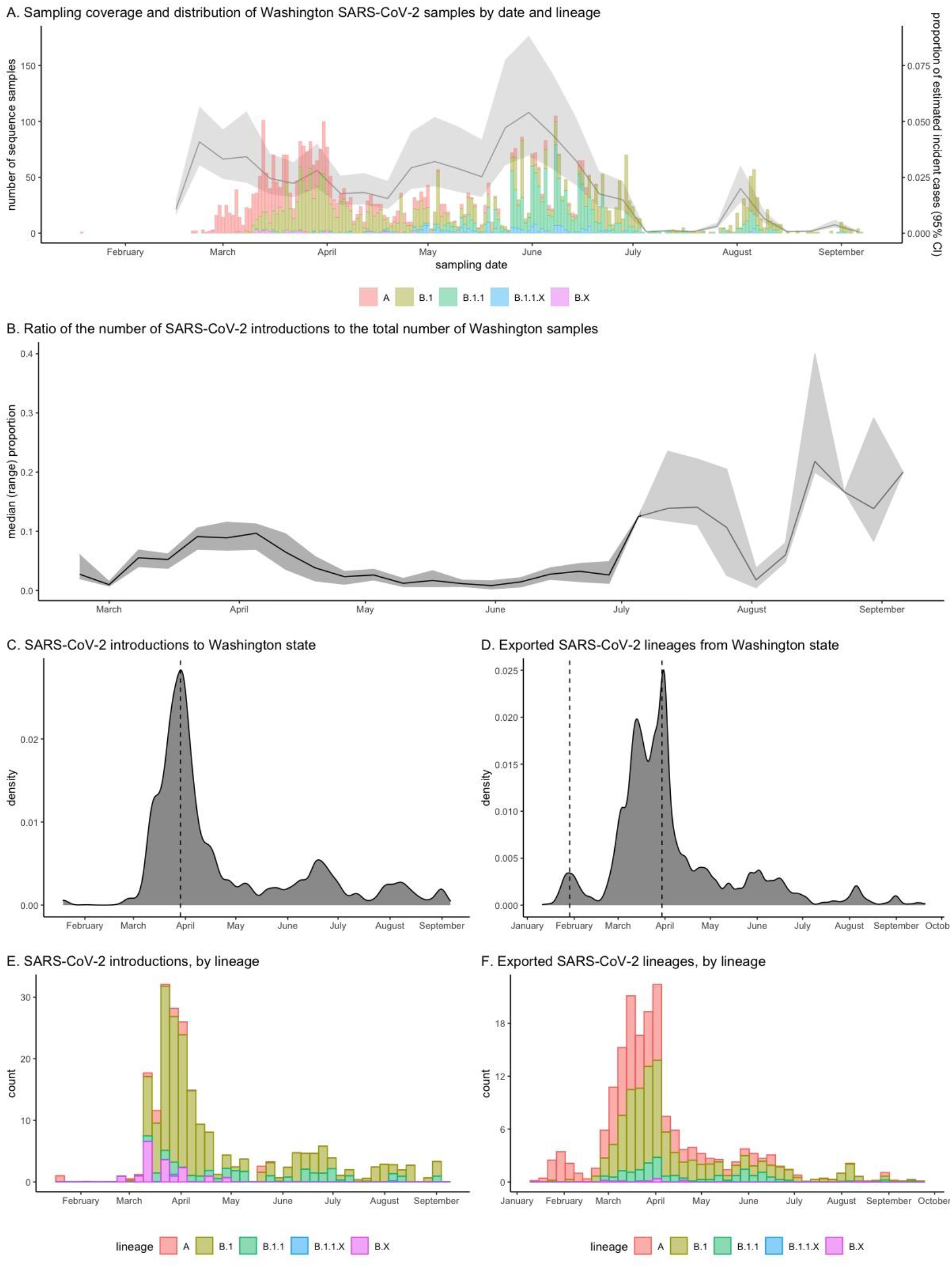
Temporal trends in SARS-CoV-2 sampling coverage, introductions and exported lineages for Washington State.

We observe temporal trends in prevalent lineage over time (Figure 3A). Early WA samples included mostly sequences of lineages A and B.1. Sequences collected in May and later were comprised largely of B.1.1 lineages. The latter of these (B.1, B.1.1, and B.1.1.X lineages) are all characterized by the Spike 614G variant. There was also substantial county-level variation in sequence sample and coverage (Figure 1B and 1C) and sampling was most representative for King and Yakima counties. Of the 2,688 sequences from western WA, 59% were sampled in King County, and of the 1,960 sequences from eastern WA, 84% were sampled in Yakima County.

We estimate that the were a minimum 287 distinct introductions (range 244-320) of SARS-CoV-2 into WA and 204 exported lineages (range 188-227) through mid-September 2020 (Figure 2). SARS-CoV-2 introductions were primarily B.1 (median 72%, range 61-77%) and B.1.1 (median 15%, range 12-20%) lineages, whereas exported lineages were primarily lineage A (median 42%, range 36-48%) and B.1 (median 47%, range 42-54%).

Most (73%) introductions occurred prior to May 1 and the number of introductions peaked on March 29, 2020, six days following WA’s “Stay-Home Stay-Healthy” order (Figure 3). When we stratify by lineage, we observe that lineage A appears to have been introduced at a small number of discrete time points (median 13, range 8-12) while the introduction of lineages B.1 and B.1.1. occur continuously over the spring and summer months. In contrast we observe two waves of exported lineages, the first peaking on January 29, 2020 and the second peaking on March 30, 2020. The first wave of exported lineages was comprised entirely of lineage A while the second wave was predominately lineages A and B.1.

Overall, the ratio of introductions to sampled sequences was 6.1% (range 5.3-6.8%), and is a proxy measure for the relative contribution of introductions to overall incidence. In our analysis, this ratio peaked in late March at 9.7% (range 7.0-11.2%) and fell to around 1-3% in May and June (Figure 3B). After July, our estimate of this ratio is less reliable due to significant under-sampling (Figure 3A).

We estimated that the majority (median 61%, range 55-65%) of introductions to WA state likely originated from a source elsewhere within the US, while the remaining 39% (range 35-45%) of introductions likely originated from outside of the US (Table 1). We also observed a significant amount of intra-state SARS-CoV-2 transmission. There were a large number of introductions from western WA into eastern WA (median 130, range 115-153), and this comprised the majority (median 65%, range 59-73%) of introductions into the eastern region on the state compared to those from elsewhere in the US (median 20%, range 12-27%) or outside of US (median 15%, range 10-21%). Conversely, there were slightly fewer distinct introductions from eastern WA into western WA (median 94, range 77-115), but these comprised a small proportion of all introductions into the western region on the state (median 28%, range 24-31%) compared to those from elsewhere in the US (median 49%, range 42-57%) or outside of US (median 23%, range 17-29%).

**Table 1.** Count and proportion of introductions into Washington state by geographic region.

| Likely Geographic | WA Overall |  | Western WA |  | Eastern WA |  |
| --- | --- | --- | --- | --- | --- | --- |
| Source of | N | % | N | % | N | % |
| Introductions | median (range) | median (range) | median (range) | median (range) | median (range) | median (range) |
| US | 177 (126-218) | 61% (51-68%) | 163 (137-191) | 49% (42-57%) | 41 (25-56) | 20% (12-27%) |
| Outside of US | 61 (52-68) | 39% (31-48%) | 77 (57-100) | 23% (17-29%) | 30 (19-41) | 15% (10-21%) |
| Intra-state |  |  |  |  |  |  |
| Western WA |  |  |  |  | 130 (115-153) | 65% (59-73%) |
| Eastern WA |  |  | 94 (77-115) | 28% (24-31%) |  |  |
| Total |  |  |  |  |  |  |

The size of WA subclades resulting from each introduction ranged from 1 to 2,193 sequences. Most introductions resulted in a single WA sequences (72%) or small subclades of 2 (9%) or 3 to 5 SARS-CoV-2 sequences (8%). The remaining 6% of introductions resulted in moderately sized subclade of 6-20 WA sequences, and 6% resulted in large subclades of 20 or more WA sequences. The duration of each subclade − defined as the number of days from first to last sequence collection − was positively correlated with subclade size (Figure 4). Subclades of just 2 sequences had the shortest duration, with a median of 5 days (IQR 1-16 days), the median duration for small subclades of 3 to 5 sequences was 28 days (IQR 11-46 days). Moderately sized and large subclades had a median duration of 38 days (IQR 20-62 days).

**Figure 4.**
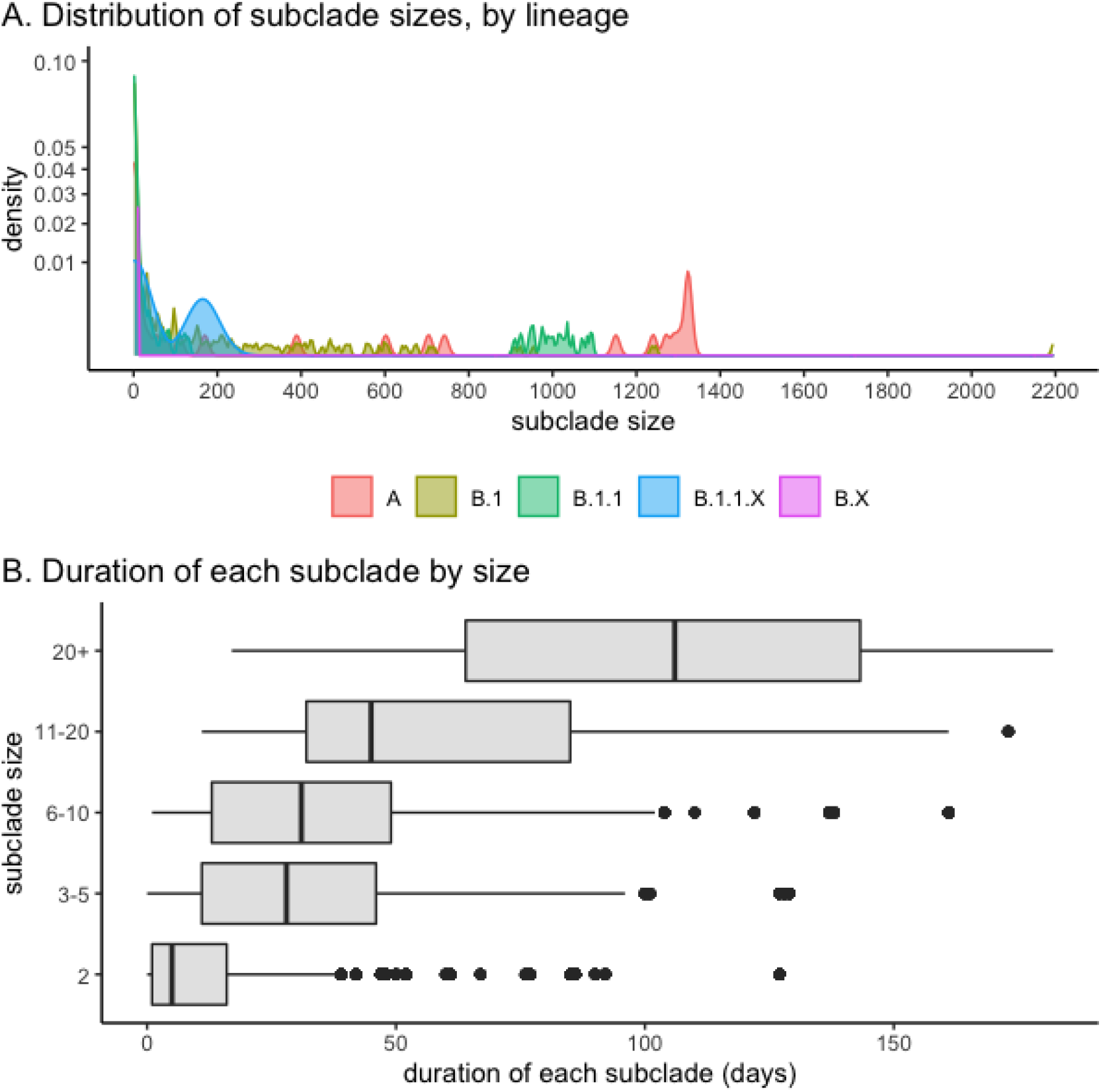
Size and duration of Washington state subclades resulting from an out-of-state introduction of SARS-CoV-2.

## DISCUSSION

We found phylogenetic evidence that the SARS-CoV-2 epidemic in WA was seeded by a large number of distinct, ongoing introductions through September 2020. We similarly estimated that a large number of SARS-CoV-2 lineages were exported from WA state during the same period. Introductions appeared to play a significant role early in the epidemic in March and April, but they continued through the summer of 2020. Notably, the peak of introductions on March 29, 2020 coincided exactly with the highest day of test-positivity in WA state.^18^ Although many introductions of SARS-CoV-2 result in a single descendent WA sequence, which is suggestive of limited local transmission, the sequence collection time frame for small clades lasted on the order of a week to one and a half months, which suggests these clades represent a modest degree of local transmission. In addition, approximately 12% of introductions resulted in larger subclades, corresponding to long chains of local SARS-CoV-2 transmission.

The majority of introductions of SARS-CoV-2 into WA likely originated from elsewhere within the US. Thus, inter-state travel within the US may play a more important role in sustaining and re-seeding local epidemics, compared to international travel. This is consistent with prior analyses which found that domestic and inter-state travel is a significant source of new SARS-CoV-2 infections, including a Connecticut outbreak that was phylogenetically linked to the initial outbreak in WA state as the likely source.^19^ We also observed a significant amount of intra-state transmission between the eastern and western regions of WA. Notably, intra-state transmission account for most transmissions into eastern WA, but the converse was not true. This asymmetrical transmission pattern may be because western WA includes infrastructure for both inter-state and international travel, including the I-5 corridor, the Seattle-Tacoma international airport, and the seaports.

We observed distinct temporal trends in introductions and exports by lineage, and lineages with the Spike 614G variant accounted for the majority (88%) of introductions. However, the temporal patterns of introductions and exported SARS-CoV-2 lineages mirrors the temporal shifts of dominant clades that occurred elsewhere in the US and globally.^8^ During the study period, none of the variants of concern from the United Kingdom (B.1.1.7 or SARS-CoV-2 VOC 202012/01), South Africa (B.1.351 or 501.V2), or Brazil (P.1) were circulating in WA. The first case of B.1.1.7 variant in WA state was sampled from Snohomish County on December 25, 2020; the first case of B.1.351 was sampled in January in King County; and the first case of P.1 was sampled in March, also in King County.

Other studies examining the origins of SARS-CoV-2 in Brazil, Northern California, New York, and Boston have also found evidence of multiple introductions.^10,20−22^ Our findings are consistent with prior analyses that quantified the relative important of introductions of SARS-CoV-2.^23,24^ Using a similar analytic approach, du Plessis et al. similarly found a large number of introduction of SARS-CoV-2 into the United Kingdom (N=1179, 95% interval 1143-1286) through June 2020; the majority of which resulted in small clades of fewer than 10 sequences.^24^ In the United Kingdom, most introductions that occurred prior to May likely originated in Italy, Spain, or France. Another analysis of the relative contribution on introductions to the SARS-CoV-2 epidemic in Switzerland found that most introduced lineages were from neighboring counties (France, Italy, Germany and Belgium).^23^ Our findings are also consistent with phylodynamic modeling during the first epidemic wave that estimated 3-10% of SARS-CoV-2 cases in WA (excluding Yakima County) were attributable to an introduction; we also observed two distinct waves of introductions, the first which included lineages with the original Spike 614D mutation (i.e. lineage A), followed by the introductions of lineages with the Spike 614G variant (i.e. lineages B.1 and B.1.1).^17^

This study has a number of strengths. To our knowledge, this is the first large-scale phylogenetic analysis to quantify the number of introductions of SARS-CoV-2 in a region of the US or to quantify within-county and within-state transmission patterns using phylogenetic methods. In addition, our findings were robust to the inclusion of different time-stratified random samples of non-WA reference sequences.

This analysis has several limitations. First, we likely are underestimating the true number of introductions due to incomplete sampling. Phylogenetic results need to be interpreted carefully due to incomplete sampling and phylogenetic uncertainty. Because our sampling coverage was only 6% of confirmed SARS-CoV-2 cases, if we were to sequence more genomes, we would find more introductions. Although GISAID and participating laboratories have facilitated the availability of an unprecedented number of publicly available whole genomes of SARS-CoV-2, within specific geographies, sampling coverage is very low and there is significant over-representation of sequences from certain countries (e.g. the United Kingdom, Australia, and the US). Second, due to length-biased sampling, we were unable to assess temporal variation in downstream clade sizes changed over time.

Our findings have several important public health implications. First, our findings highlight the importance of genomic surveillance to monitor for emerging variants in WA and elsewhere in the US due to the high levels of inter- and intra-state transmission of SARS-CoV-2 lineages.^25^ Monitoring inter- and intra-state transmissions and their origins can be used to determine where public health interventions may be most effective. In addition, we observed that number of both introductions and exports fell within the week of the “Stay Home, Stay Health” order, suggesting that lockdowns may be effective at immediately reducing inter- and intra-state transmission.

## Data Availability

The data that support the findings of this study are openly available via GISAID at https://www.gisaid.org/.

## ACKNOWLEDGEMENTS

We would like to acknowledge Dr. Niket Thakker for providing incidence estimates and reviewing the manuscript, as well as the UW Virology staff who assisted to sequence collection and assembly. We gratefully acknowledge the originating laboratories responsible for obtaining the specimens and the submitting laboratories where genetic sequence data were generated and shared via the GISAID Initiative, on which this research is based. DMT receives support from the NIH National Institute of Allergy And Infectious Diseases (F31AI152542). JTH and DMT also receive support from the NIH National Institute of Allergy and Infectious Disease (R01AI127232). Funders had no role in study design, collection, analysis, and interpretation of data, writing of the report, or the decision to submit the paper for publication.

## CONTRIBUTIONS

DMT and JTH contributed to the study conceptualization, methodology, writing of the original draft, and accessed and verified the data. DMT conducted the formal analysis, visualization. JTH contributed to supervision. ALG, PR, LS, HX, KRJ, NB, and MLH collected and analysed laboratory data (including genomic sequencing) and contributed to data curation. MF contributed to methodology and interpreted results. All authors reviewed, edited, revised and gave final approval of the Article before submission.

